# Risk of extrapyramidal side effects in the newborn after single dose maternal metoclopramide administration prior to cesarean section: an analysis via physiology-based pharmacokinetic and pharmacodynamic modeling

**DOI:** 10.64898/2026.10.08.26365051

**Authors:** Hedwig van Hove, Joyce E.M. van der Heijden, Petra van den Broek, Joris van Drongelen, Saskia N. de Wildt, Rick Greupink

## Abstract

**Background:** Metoclopramide is prescribed during pregnancy to treat nausea and vomiting. Despite its widespread application, there is potential for peripartum maternal metoclopramide dosing to induce extrapyramidal symptoms (EPS) in the newborn due to residual pediatric drug levels at birth. Here, a physiology-based pharmacokinetic model, including pregnancy-related changes and placental transfer values, was established and verified for metoclopramide. Human placenta cotyledon perfusion experiments were conducted to estimate placental transfer of metoclopramide. Transfer parameters were incorporated in a Simcyp PBPK model to enable predictions of fetal plasma and brain concentrations at different gestational ages (GA). Simulations were performed for the pre-cesarian section dosing of 10 milligram (mg) intravenously (IV, single dose), after which predicted fetal brain concentrations were used to calculate dopamine (D_2_) receptor occupancy.

**Findings & Conclusions:** Human *ex vivo* placental perfusion data were successfully generated and integrated into a pregnancy-specific physiology-based pharmacokinetic model to characterize maternal, fetal, and fetal brain exposure to metoclopramide. The model adequately predicted pharmacokinetics in healthy volunteers and pregnant individuals. Simulations of 10 mg IV dosing show mean metoclopramide peak concentrations of 47.0, 39.5 and 37.9 ng/mL in the fetal plasma at 24, 32 and 40 weeks GA, respectively. Residual drug levels immediately after birth were assumed to be equal to predicted fetal plasma levels. Newborn brain concentrations were predicted to be of the same order of magnitude as maternal plasma levels, with maternal plasma-to-fetal brain ratios of 1:0.98, 1:0.76 and 1:0.65 at GA 24, 32 and 40 weeks, highlighting evident distribution to the newborn central nervous system. Average newborn brain D_2_ receptor occupancy, which was used as a surrogate pharmacodynamic marker for EPS, remained below the 80% threshold for all simulated GAs. However, predicted population variability indicated that for a small but clinically relevant subset of newborns (∼ 5% at 24 weeks and ∼1% at 32 weeks GA, respectively), the 80% D_2_-receptor occupancy could be exceeded under standard dosing regimens, indicative of EPS risk.

## INTRODUCTION

The combined dopamine and serotonin receptor antagonist metoclopramide is approved by the Food and Drug Administration (FDA) and different national evaluation boards in the EU to treat nausea and vomiting in patients with gastroesophageal reflux disease or diabetic gastroparesis by increasing gastric motility. It is allowed to prescribe short-term during pregnancy to treat general nausea and vomiting, as well as hyperemesis gravidarum. About 50% of women experience nausea and vomiting during their pregnancy, and about 10% of them receives pharmacological intervention (1). Metoclopramide is often recommended, but despite its widespread application in treating pregnancy-associated nausea and vomiting, the safety profile of metoclopramide in pregnancy remains a subject of ongoing research and debate (2).

Specifically in the case of cesarean section, metoclopramide is one of the treatment options used preoperatively, primarily for its antiemetic effects to prevent nausea and vomiting, which are frequent complications in this patient population. Mishriky et al. demonstrate that the prokinetic action of metoclopramide enhances gastric emptying and promotes gastrointestinal motility, thereby significantly reducing the incidence of perioperative nausea and vomiting. This results in the primary clinical benefit of metoclopramide when administered prior to cesarean delivery: as an effective prophylactic agent against nausea and vomiting in cesarean sections, especially in patients at increased risk for these complications (3).

Although metoclopramide seems to be effective, concerns have been raised regarding its potential to cause extrapyramidal symptoms (EPS) in the newborn. EPS can manifest as abnormal muscle tone (either increased stiffness or hypotonia), tremors or shaking, involuntary movements, such as dystonia or difficulty feeding due to impaired motor control. Next to this, EPS have also been reported with greater frequency in children compared with adults when receiving metoclopramide (4, 5).

Metoclopramide can cross the placenta and the blood-brain barrier (BBB) and may theoretically affect the fetal central nervous system, raising the possibility of neonatal EPS (6). Supporting this, a pharmacovigilance review reported rare but documented cases of neonatal EPS following maternal metoclopramide exposure during pregnancy, highlighting the need for cautious use (7). Notably, the European Medicines Agency (EMA) contraindicates the use of metoclopramide in children under 1 year of age due to concerns for EPS (5).

Ideally, since it is ethically challenging to conduct clinical studies in pregnant patients, existing data should be used as effectively as possible. Unfortunately, even for commonly prescribed medications like metoclopramide, data on safety and exposure during pregnancy are scarce. This makes *in vivo* benefit-risk assessments and determining an optimal dosing strategy, challenging for pregnant women and their fetuses. Physiology based pharmacokinetic (PBPK) modeling is an *in silico* technique that enables internal exposure predictions. The models incorporate physiological changes that take place during gestation, this way they can help refine dosing recommendations by simulating various dosing scenarios and predicting drug concentrations in both the mother and the unborn child (8).

Our research aims to assess maternal and fetal metoclopramide exposure after pre–cesarean section dosing using PBPK modeling and simulation. Next, we aim to link fetal exposure to dopamine receptor occupancy as a marker for risk of fetal EPS.

## MATERIALS AND METHODS

Briefly, in this study we first developed a metoclopramide PBPK model, which was extended with experimentally determined placental transfer parameters and was also adapted to capture maternal and fetal physiological changes that occur during pregnancy. The metoclopramide compound model was newly built, the population models describing physiology were directly available in the Simcyp software. Model performance was first verified against available clinical data before current metoclopramide dosing regimens were simulated to assess fetal plasma and brain concentrations. With these outcomes, dopamine receptor occupancy in the newborn was calculated, based on D_2_ receptor affinity, and predicted unbound brain concentrations, using a simple B_max_ model, i.e. according to the Hill equation (9). Outcomes were compared to the threshold for EPS risk, which correlates well with D_2_ receptor occupancy and has been reported at 80% receptor occupancy based on studies with a wide range of clinically applied D_2_ receptor antagonists, for example drugs used for psychiatric indications (10).

### 2.1 Human placenta perfusion experiments

Placentas from uncomplicated (late-)third trimester pregnancies were obtained from consenting women who underwent elective caesarean sections or vaginal delivery. For this, approval of the institutional medical ethical committee was obtained (CMO Arnhem/Nijmegen File 2014–1397 and 2022-13523) in line with The Code of Ethics of the World Medical Association (Declaration of Helsinki).

To study the transplacental transfer of metoclopramide, *ex vivo* dual-side placenta perfusions were performed in a closed-closed experimental configuration as reported previously by van der Heijden et al. (11). Metoclopramide concentrations in perfusates were analyzed via LC-MS/MS. For quality control purposes, antipyrine was included in the perfusions as passive diffusion marker. Additionally, unwanted leakage of perfusion buffer from the circulations was monitored throughout the 180-minute study. A more detailed description on the methodologies can be found in the Supplementary Information.

The data obtained from the placental perfusion experiments were subsequently analyzed with a physiologically-based semi-mechanistic placenta (PBMP) model to derive the intrinsic unbound transport clearance values that describe the transport of metoclopramide between the different placental compartments in the ex vivo system (12). The unbound intrinsic transport clearance values that followed from this analysis were then incorporated in the Simcyp pregnancy PBPK model to enable predictions of maternal and fetal pharmacokinetics.

### 2.2 Healthy volunteer and pregnancy PBPK models

All simulations were conducted using Simcyp v21 (Certara UK Limited, Simcyp Division, Sheffield, UK). Before simulations could start, a compound model for metoclopramide was established, based on data collated from literature, see Supplementary Information, Table S2. For all simulations, the default physiology profiles for healthy volunteers, pregnant women or fetus in this version of Simcyp were used (13). To verify the model against clinical data, healthy non-pregnant volunteer as well as pregnancy pharmacokinetic (PK) data for metoclopramide were mined from PubMed (Table S3 and S4). For a visual check of predicted versus observed concentration-time (Ct) profiles, observed plasma Ct profiles were extracted from literature and digitalized with WebPlotDigitizer v4.6. For this work, the proposed workflow as described by van der Heijden et al. was applied (14). Predictive performance of the healthy volunteer and pregnant women PBPK model was evaluated by a visual predictive check and by calculating the ratio of predicted-to-observed (p/o) PK parameters. P/o ratios within 0.5 to 2-fold range were considered acceptable (15).

To predict fetal exposure, the placenta perfusion transfer values obtained in the *ex vivo* experiments were included in the PBPK model to simulate placental transfer. To assess whether gestational changes influence metoclopramide exposure during pregnancy, simulations for different relevant gestational age (GA) groups were performed (i.e., GA of 24, 32 and 40 weeks). A single dose of 10 mg intravenous (IV) was used to predict maternal and fetal metoclopramide plasma concentrations and fetal brain concentrations in late-second and third trimester (GA 24, 32 and 40 weeks).

### 2.3 Dopamine receptor occupancy of metoclopramide

Predicted fetal plasma concentrations of metoclopramide following dosing of a single dose of 10 mg IV were used to predict fetal brain exposure using a Simcyp-predicted brain-to-plasma partition coefficient specific to the fetal compartment. This partition coefficient was established by the Rodgers and Rowland method from physicochemical properties of metoclopramide (being mainly the acid dissociation constant (pKa), lipophilicity, and fraction unbound) (16, 17). Subsequently, receptor occupancy of fetal dopamine D₂ receptors was calculated using the Hill equation, incorporating estimated unbound fetal brain concentrations (unbound fraction of 0.295, (18)) and published affinity (KA) values for metoclopramide:

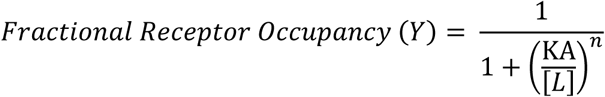

In which L represents the concentration of the ligand, in this case metoclopramide, KA is the ligand concentration producing 50% receptor occupancy, and n the Hill coefficient, which reflects the degree of cooperativity in ligand binding, respectively. Values for L were predicted using the PBPK model, KA was reported in literature as 28.8 nM (19), and n was reported to be 1 (20). Predicted receptor occupancy levels were then compared to an established threshold value of 80% D_2_ receptor occupancy, above which the risk of EPS is considered to be elevated (10).

## RESULTS

### 3.1 Placental passage metoclopramide

Three successful placenta perfusions with metoclopramide were performed in cotyledons obtained from different donors (Supplementary Information, Table S1). In all experiments volume leakage from the fetal to maternal buffer was minimal at the end of the perfusions (< 3 mL/h). In addition, antipyrine transport reached equilibrium after 120 minutes of perfusion, indicating successful cannulation of the cotyledon, i.e., overlap of the maternal and fetal circuits (Supplementary Information, Figure S1). Metoclopramide showed extensive transport over the placental barrier, see Figure 1.

**Figure 1.**
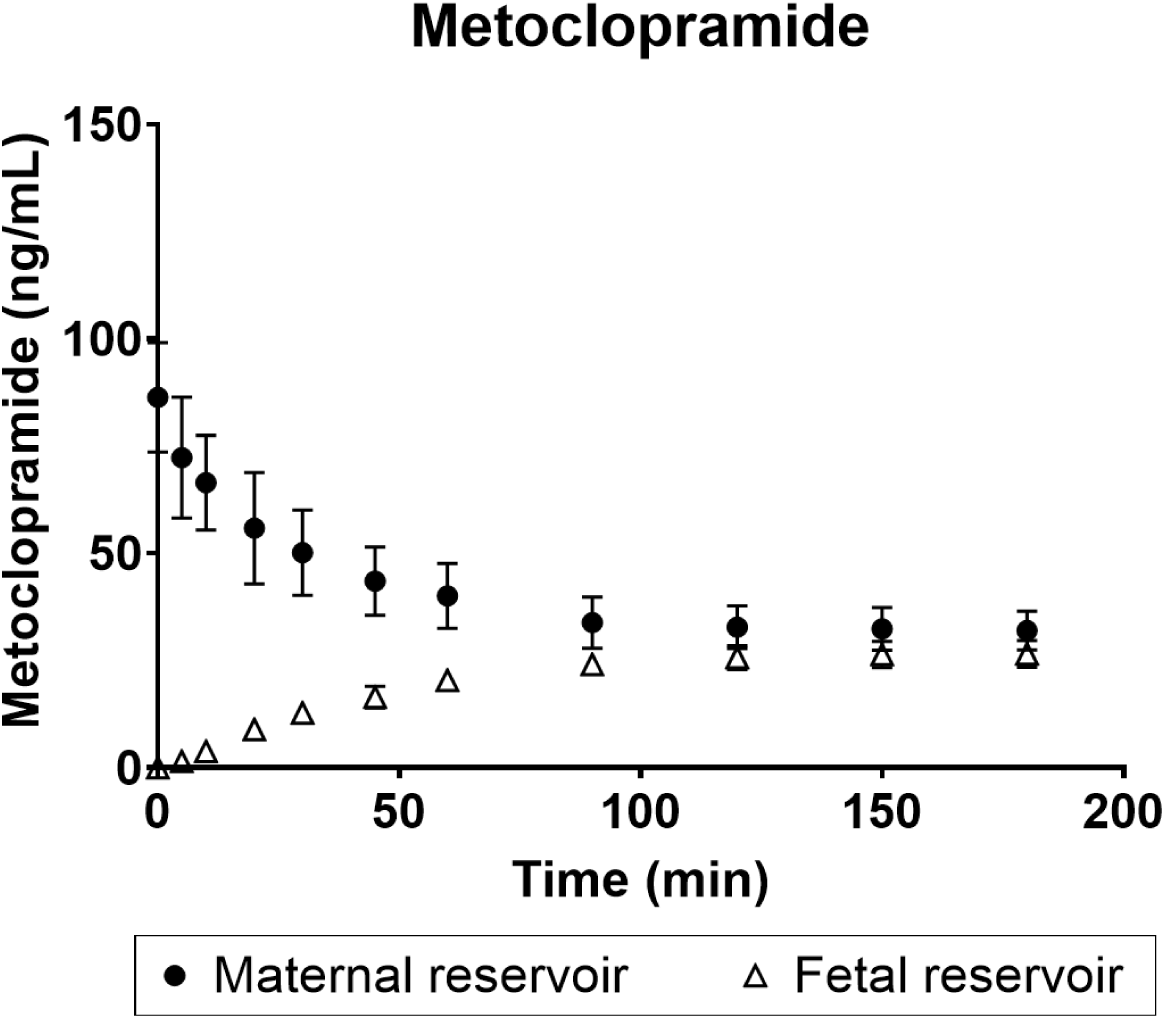
Course of metoclopramide concentration over time during human *ex vivo* placenta perfusions. (**21**)

The data obtained from the placental perfusion experiments (Figure 1) were subsequently used in the PBPM model to derive the intrinsic unbound transport clearance values that describe the transport of metoclopramide between the different placental compartments (12, 22). These outcomes were compatible with the Simcyp-based PBPK model, see Supplementary Information Table S2 (Placental Intrinsic unbound clearance values). Mean values were predicted to be 683.9, 420.6, 2360.8 and 1198.5 µL/min/mL placenta for maternal circulation-to-placenta, placenta-to-maternal circulation, fetal circulation-to-placenta and placenta-to-fetal circulation, respectively, and were incorporated into the PBPK model to simulate fetal exposure.

### 3.2 Predictive performance PBPK models

#### Healthy volunteers

Overall, the PBPK model was considered adequate to predict PK in healthy volunteers, since after a single IV dose, the mean ± SD p/o ratio was 1.13 ± 0.13 for the volume of distribution, 1.48 ±0.2 for the clearance, 1.72 ± 0.03 for the maximal concentration (C_max_), 0.94 ± 0.25 for the time to maximal concentration (t_max_), 1.22 ± 0.54 for the half-life (t_1/2_) and 0.83 ± 0.56 for the area under the curve (AUC), respectively. End after a single PO administration, the average ± SD p/o ratio was 0.96 ± 0.32 for the C_max_, 0.78 ± 0.23 for the t_max_, 1.02 ± 0.29 for the t_1/2_ and 0.58 ± 0.20 for the AUC, respectively (Supplementary Information, Figure S3). Visual predictive checks were conducted for IV (Supplementary Information, Figure S4) or PO administration (Supplementary Information, Figure S5) in healthy volunteers and deemed adequate. Observed data were obtained from different pharmacokinetic studies (23–37).

#### Pregnant individuals

Simulated plasma Ct profiles of pregnant women receiving a 10, 14 or 15 mg/kg IV dose were predicted reasonably well, yet appear to be slightly underestimated (Figure 2A, C, E). Fetal exposure after dosing was predicted with similar accuracy (Figure 2B, D and F). Since no PK parameters was reported in literature, no p/o ratios could be determined. Taking all together, model performance was considered adequate to predict PK in pregnant women and their newborns.

**Figure 2.**
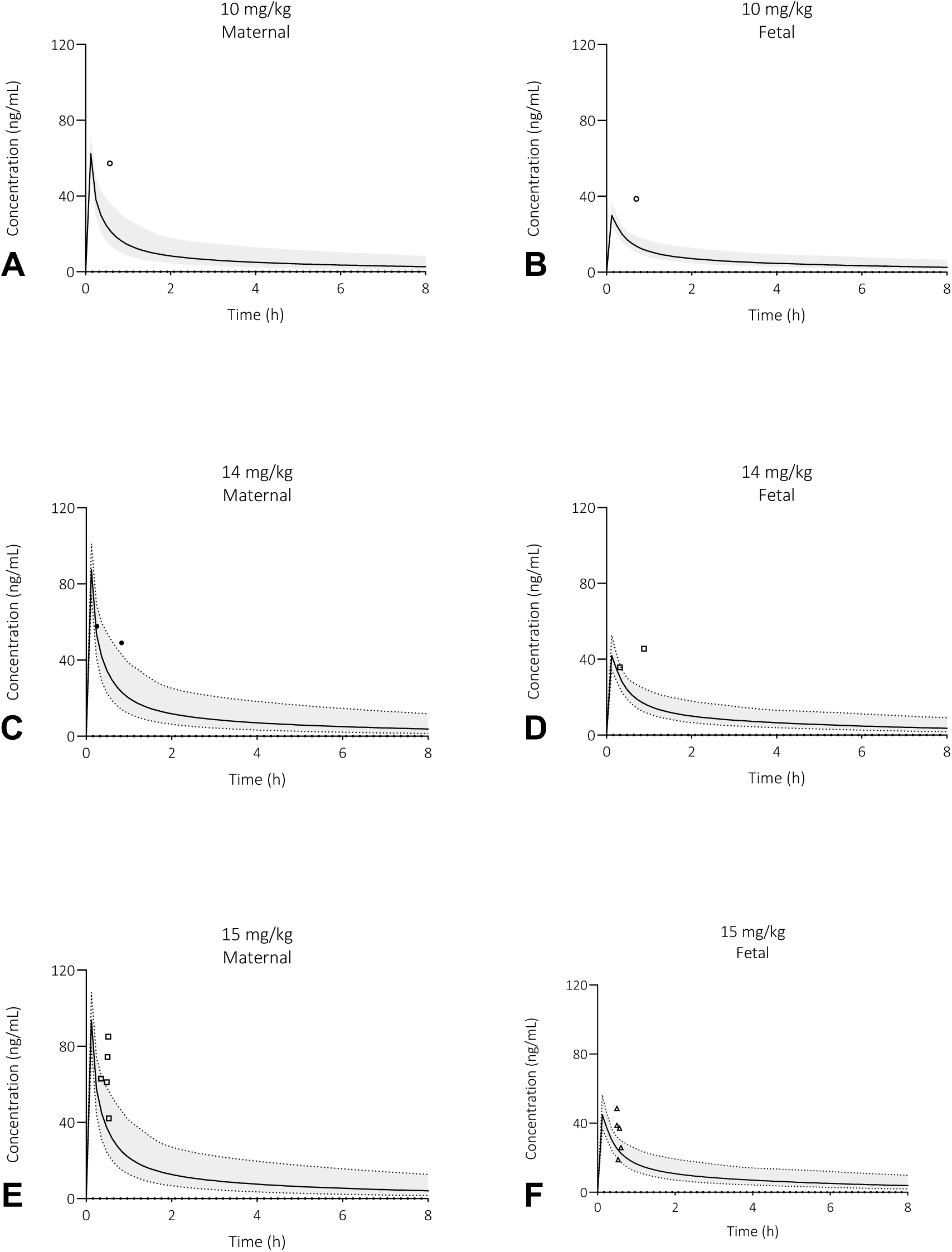
Prediction of metoclopramide plasma concentration-time profiles in pregnant women and their fetuses after a single dose intravenous (IV) administration. The solid line is the predicted mean of the simulated population and the shaded area represents the 5^th^ to 95^th^ percentile of the virtual population. Closed circles are the observed individual data points after a 10 (A,B), 14 (C,D) or 15 (E,F) mg/kg IV dose (6). Abbreviations: milligram per kilogram bodyweight (mg/kg).

### 3.3 Current dosing regimen in relation to D_2_ receptor occupancy in newborns

Following the administration of metoclopramide at 10 mg IV single dose, the PBPK model predicts that mean fetal plasma concentrations reach approximately 50% of mean maternal plasma levels at all GAs (Figure 3). Notably, mean fetal brain concentrations equal those observed in maternal plasma, indicating preferential distribution or accumulation in fetal brain tissue. A considerable interindividual variability is observed in predicted fetal brain exposure, with a wide range between GAs. Since our focus is assessing exposure after dosing immediately prior to cesarean section, we assume that residual fetal drug levels at birth will closely approximate fetal exposure in scenarios modeled at 24, 32 or 40 weeks of gestation, depending of their GA at birth.

**Figure 3.**
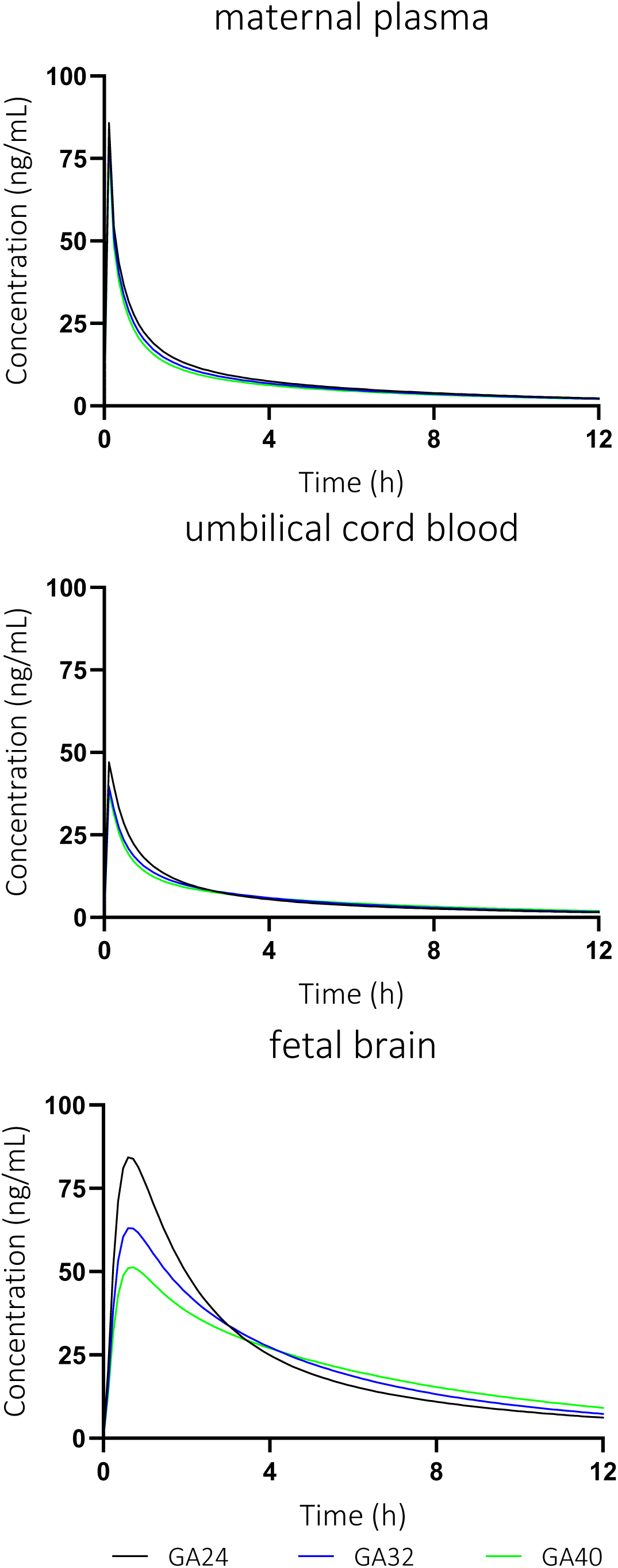
Prediction of mean total metoclopramide concentration in maternal plasma, umbilical cord and in fetal brain after a single 10 mg IV dose. Virtual population consisted of 100 subjects at gestational ages of 24, 32 or 40 weeks, respectively.

Based on the predicted fetal plasma concentrations, we estimated that the mean dopamine D_2_-receptor occupancy in the newborn brain was below the EPS threshold of 80% (Figure 4). The majority of simulated newborns fell well under this benchmark. However, a subset of the population (approximately 5%) exceeded the 80% occupancy threshold at GA 24 weeks. For 32- and 40-weeks GA, predictions show a decreased risk in EPS development, with ∼1% exceeding the threshold at GA 32 weeks, and no predicted individuals exceeding the threshold at 40 weeks GA, respectively (Figure 5). This finding suggests that metoclopramide is likely safe for most newborns, but a small proportion of newborns may be at risk for EPS-related effects under the current dosing regimen.

**Figure 4.**
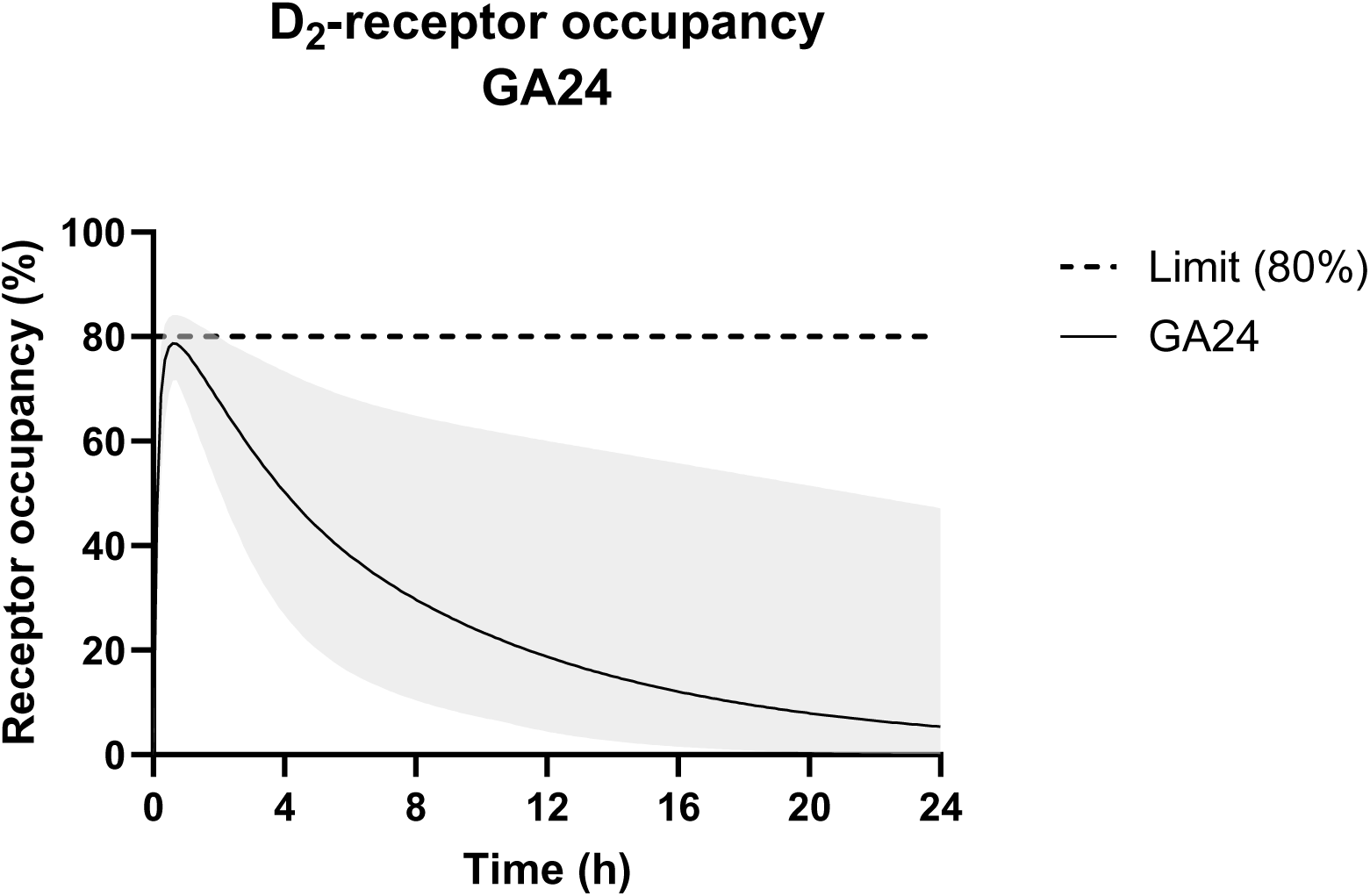
Prediction of newborn brain D_2_-receptor occupancy 10 mg IV dosing of the mother at GA 24 weeks. The solid lines are the predicted arithmetic means and the shaded area represents the complete population range. The dashed line represents the threshold value associated with extrapyramidal side effects risk, as assessed in adults for a range of D_2_-receptor antagonists (10), we also assumed relevant for the newborn population (80%). Virtual population consisted of 100 subjects with a gestational age of 24 weeks.

**Figure 5.**
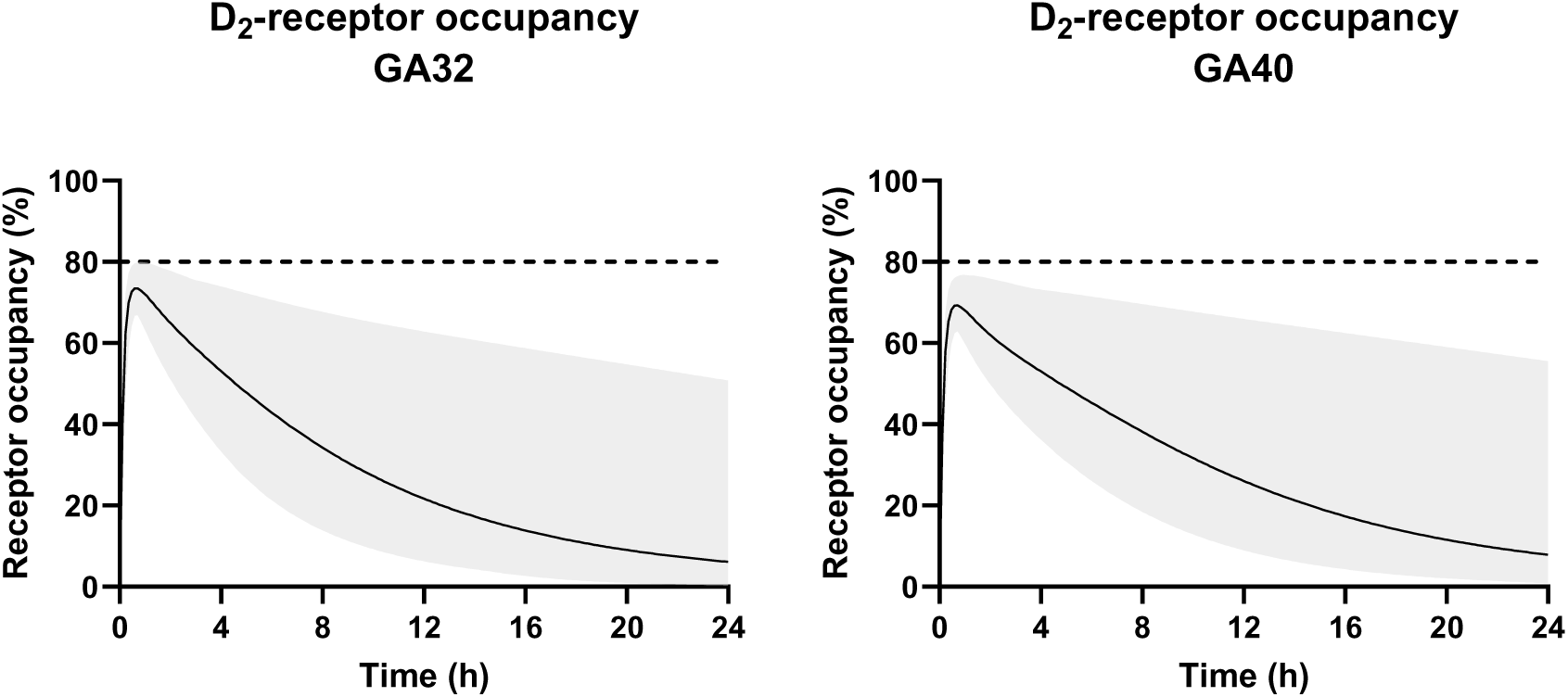
Prediction of newborn brain D_2_-receptor occupancy after 10 mg IV dosing of the mother (GA32 + 40). The solid lines are the predicted arithmetic means and the shaded area represents the complete population range. The dashed line represents the threshold value associated with extrapyramidal side effects risk, as assessed in adults for a range of D_2_-receptor antagonists (10), we also assumed relevant for the newborn population (80%). Virtual population consisted of 100 subjects with gestational ages of 32 or 40 weeks, respectively.

## DISCUSSION

In this study, we combined *ex vivo* human placental perfusion data with a pregnancy-specific PBPK modeling framework to predict fetal exposure and pharmacodynamic outcomes of metoclopramide following standard maternal dosing. By integrating experimentally derived placental transfer parameters into the PBPK model, we were able to simulate not only maternal and fetal plasma concentrations but also fetal brain exposure and downstream dopamine D_2_ receptor occupancy, as a pharmacodynamic surrogate endpoint for the risk of neonatal EPS.

A key strength of this study is the mechanistic incorporation of human-derived placental transfer data, which enhances the biological relevance of fetal exposure predictions. This integration addresses a common limitation in pregnancy PBPK modeling, where transplacental kinetics are often assumed or inferred from limited clinical data (38). Furthermore, the use of established receptor binding kinetics for metoclopramide on the D_2_ receptor allowed us to move beyond exposure assessment and quantitatively estimate fetal receptor occupancy, a more pharmacodynamically meaningful outcome which stands in relation to drug effects. Particularly D_2_ receptor occupancy, more precisely D_2_ receptor antagonism, as for example assessed for antipsychotics, holds a tight relation to EPS. To our knowledge, this is the first study to translate predicted fetal metoclopramide brain exposure to predicted dopamine D_2_ receptor occupancy in utero / right after birth. Incorporating fetal or newborn pharmacodynamics into perinatal drug evaluation represents a critical advancement toward improving drug safety and optimizing dosing strategies during pregnancy.

Our results show that while average fetal D_2_ receptor occupancy remains below the 80% threshold associated with EPS, a proportion (8.1%) of the virtual newborn population exceeds this benchmark. These findings suggest that, under standard dosing conditions, a subset of newborns may be at increased risk for EPS. It should be noted that the proportion of newborns exceeding the 80% receptor occupancy threshold may be influenced by factors such as the timing of the last maternal dose relative to delivery, which was not accounted for in our simulations since steady-state was simulated. Thus, the current predictions represent a worst-case scenario that likely (slightly) overestimates the actual clinical risk. Still, our findings support that a cautious approach to metoclopramide use during pregnancy, especially in repeated or high-dose regimens close to delivery, should be considered.

A major limitation of the study is that, while receptor occupancy provides a useful surrogate for pharmacodynamic effects, the translation of occupancy thresholds is derived from adult data. It is uncertain whether this threshold is accurate for newborns or children and therefore needs more investigation. Neonatal sensitivity to D_2_ antagonism may differ due to developmental differences in receptor density or neurochemical environment. This concern is further supported by existing clinical guidelines of the European Medicines Agency (EMA) and the US Food and Drug Administration (FDA), which are against the use of metoclopramide in children under one year of age due to their increased sensitivity to extrapyramidal symptoms, which is a vulnerability that may similarly apply to the fetus and newborn (39).

Although Simcyp version 21 allows prediction of fetal brain drug concentrations, it does not currently account for all unique features of the developing BBB. Specifically, the model lacks detailed information on developmental differences in BBB structure and permeability, such as the ontogeny of tight junctions, efflux transporters (e.g., P-gp, BCRP), and pericyte coverage (40, 41). Current simulations of brain concentrations use partition coefficients based on the method by Rodgers and Rowland to predict partition coefficients for drug tissue distribution (16). This method is mechanistic and accounts for drug ionization. LogP and fraction unbound in plasma are the main contributors to brain exposure, and are well-established drivers of tissue distribution (17). However, it assumes passive, non-saturable distribution, making it suboptimal for certain drug classes (for example in the case of drugs that are subject to active transport) (16, 17). Although the method is expected to be suitable for metoclopramide, in theory the model would benefit from a more mechanistic and physiological approach, incorporating more features specific for the fetal BBB. It is reassuring that many key mechanisms, including functionally effective tight junctions, are already present in the newborn brain. This supports the view that the fetal and neonatal BBB is not as permissive or “leaky” as often assumed (42, 43). Indeed, studies demonstrate active transport across the fetal BBB from early developmental stages, including essential substrates like glucose and amino acids (44, 45). Nonetheless, important physiological differences remain. For example, rising systemic blood pressure and increasing plasma protein concentrations during gestation influence hydrostatic and colloid osmotic pressures, potentially affecting fluid exchange across the capillary endothelium (42).

In addition, the model software partially accounts for brain tissue maturation effects that could alter drug partitioning or clearance into fetal brain compartments. It includes the increase in brain tissue volume over gestation, blood flow alterations over gestation in the brain, and tissue composition (lipids, intra- and extracellular water). Therefore, although the model is not yet comprehensive, we believe it currently provides the best available *in silico* estimation of fetal brain exposure.

Clinically, these findings offer valuable insights into the risk-benefit balance of metoclopramide use in pregnant patients, particularly when used perioperatively for cesarean section at lower gestational ages (32 weeks and prior). While the drug remains effective for gastrointestinal symptom control, its potential impact on the developing fetal CNS must be carefully weighed, especially in high-risk populations or repeated-dose scenarios. Next to this, our work highlights the potential utility of PBPK-PD modeling in refining dosing recommendations and guiding clinical decision-making in pregnancy, where empirical data are often lacking. Our findings underscore the need for individualized risk assessment in the pharmacological management of pregnancy-related conditions and support the ongoing development of model-informed precision dosing strategies for maternal-fetal medicine. Future work should assess the impact of gestational age on fetal CNS exposure, as differences in the BBB across developmental stages may further influence the extent to which drugs cross into the fetal brain and potentially cause harm.

## ACKNOWLEDGEMENTS

We thank Marlies Peters-Knuiman (Radboudumc) for the work related to the inclusion of pregnant women. We also thank Dr. Ping Zhao (Foundation) for useful discussions that improved the manuscript. Last, we want to thank Anne van Uden and Emmely Wagener for their contributions to the experimental aspects of the research.

## REFERENCES

1. Niebyl JR. Nausea and vomiting in pregnancy. New England Journal of Medicine. 2010;363(16):1544–50.

2. Andrade SE, Gurwitz JH, Davis RL, Chan KA, Finkelstein JA, Fortman K, et al. Prescription drug use in pregnancy. American journal of obstetrics and gynecology. 2004;191(2):398–407.

3. Mishriky B, Habib A. Metoclopramide for nausea and vomiting prophylaxis during and after Caesarean delivery: a systematic review and meta-analysis. British journal of anaesthesia. 2012;108(3):374–83.

4. (FDA) UFaDA. REGLAN Injection Package Insert (metoclopramide injection, USP) 2010 [Available from: https://www.accessdata.fda.gov/drugsatfda_docs/label/2010/017862s063lbl.pdf.

5. (EMA) EMA. European Medicines Agency recommends changes to the use of metoclopramide. 2013.

6. Bylsma-Howell M, Riggs K, McMorland G, Rurak D, Ongley R, McErlane B, et al. Placental transport of metoclopramide: assessment of maternal and neonatal effects. Canadian Anaesthetists’ Society Journal. 1983;30:487–92.

7. Lau Moon Lin M, Robinson PD, Flank J, Sung L, Dupuis LL. The safety of metoclopramide in children: a systematic review and meta-analysis. Drug safety. 2016;39:675–87.

8. Thépaut E, Brochot C, Chardon K, Personne S, Zeman F. Pregnancy-PBPK models: how are biochemical and physiological processes integrated? Computational toxicology. 2023;27:100282.

9. Wagner J. Kinetics of pharmacologic response I. Proposed relationships between response and drug concentration in the intact animal and man. Journal of Theoretical Biology. 1968;20(2):173–201.

10. Kapur S, Zipursky R, Jones C, Remington G, Houle S. Relationship between dopamine D2 occupancy, clinical response, and side effects: a double-blind PET study of first-episode schizophrenia. American journal of psychiatry. 2000;157(4):514–20.

11. Van Der Heijden JE, Van Hove H, Van Elst NM, Van Den Broek P, Van Drongelen J, Scheepers HC, et al. Optimization of the betamethasone and dexamethasone dosing regimen during pregnancy: a combined placenta perfusion and pregnancy physiologically based pharmacokinetic modeling approach. American Journal of Obstetrics and Gynecology. 2025;232(2):228. e1-. e9.

12. Roelofsen D, van Hove H, Bukkems V, Russel F, Eliesen G, Greupink R. Predicting fetal exposure of crizotinib during pregnancy: Combining human ex vivo placenta perfusion data with physiologically-based pharmacokinetic modeling. Toxicology in Vitro. 2022;85:105471.

13. Abduljalil K, Furness P, Johnson TN, Rostami-Hodjegan A, Soltani H. Anatomical, physiological and metabolic changes with gestational age during normal pregnancy: a database for parameters required in physiologically based pharmacokinetic modelling. Clinical pharmacokinetics. 2012;51:365–96.

14. van der Heijden JE, Freriksen JJ, de Hoop-Sommen MA, Greupink R, de Wildt SN. Physiologically-Based Pharmacokinetic Modeling for Drug Dosing in Pediatric Patients: A Tutorial for a Pragmatic Approach in Clinical Care. Clinical Pharmacology & Therapeutics. 2023;114(5):960–71.

15. Kuemmel C, Yang Y, Zhang X, Florian J, Zhu H, Tegenge M, et al. Consideration of a credibility assessment framework in model-informed drug development: potential application to physiologically-based pharmacokinetic modeling and simulation. CPT: pharmacometrics & systems pharmacology. 2020;9(1):21–8.

16. Rodgers T, Rowland M. Physiologically based pharmacokinetic modelling 2: predicting the tissue distribution of acids, very weak bases, neutrals and zwitterions. Journal of pharmaceutical sciences. 2006;95(6):1238–57.

17. Yau E, Olivares-Morales A, Gertz M, Parrott N, Darwich AS, Aarons L, et al. Global sensitivity analysis of the Rodgers and Rowland model for prediction of tissue: plasma partitioning coefficients: assessment of the key physiological and physicochemical factors that determine small-molecule tissue distribution. The AAPS journal. 2020;22(2):41.

18. Di L, Umland JP, Chang G, Huang Y, Lin Z, Scott DO, et al. Species independence in brain tissue binding using brain homogenates. Drug metabolism and disposition. 2011;39(7):1270–7.

19. Matsui A, Matsuo H, Takanaga H, Sasaki S, Maeda M, Sawada Y. Prediction of catalepsies induced by amiodarone, aprindine and procaine: similarity in conformation of diethylaminoethyl side chain. The Journal of pharmacology and experimental therapeutics. 1998;287(2):725–32.

20. Walkembach J, Brüss M, Urban BW, Barann M. Interactions of metoclopramide and ergotamine with human 5-HT3A receptors and human 5-HT reuptake carriers. British journal of pharmacology. 2005;146(4):543–52.

21. Eliesen GA, Fransen M, van Hove H, van den Broek PH, Greupink R. Placental transfer of tofacitinib in the ex vivo dual-side human placenta perfusion model. Current Research in Toxicology. 2024;6:100149.

22. Bukkems VE, van Hove H, Roelofsen D, Freriksen J, van Ewijk-Beneken Kolmer E, Burger D, et al. Prediction of maternal and fetal doravirine exposure by integrating physiologically based pharmacokinetic modeling and human placenta perfusion experiments. Clinical Pharmacokinetics. 2022;61(8):1129–41.

23. Scaglione F, Scanni A, Tomirotti M, Dimaiuta M, Ferrari P, Fraschini F. Pharmacokinetics and bioavailability of metoclopramide nasal spray versus metoclopramide intravenous in healthy volunteers and cancer patients. Arzneimittel-forschung. 1993;43(9):986–8.

24. Ward M, Buss D, Ellershaw J, Nash A, Routledge P. Bioavailability of intranasal metoclopramide. British journal of clinical pharmacology. 1989;28(5):616–8.

25. Rotmensch HH, Mould GP, Sutton JA, Kilminster S, Moller C, Pero RW. Comparative Central Nervous System Effects and Pharmacokinetics of Neu-metoclopramide and Metoclopramide in Healthy Volunteers. The Journal of Clinical Pharmacology. 1997;37(3):222–8.

26. Graffner C, Lagerstrom P, Lundborg P, Ronn O. Pharmacokinetics of metoclopramide intravenously and orally determined by liquid chromatography. British journal of clinical pharmacology. 1979;8(5):469–74.

27. Tzachev C, Popov T. Comparative bioavailability of metoclopramide hydrochloride at a dose of 10 mg, administered in tablet form orally, intravenously and through a nasal mucoadhesive spray. J Clin Med. 2010;3(1):31–8.

28. Bateman D, Kahn C, Davies D. The pharmacokinetics of metoclopramide in man with observations in the dog. British journal of clinical pharmacology. 1980;9(4):371–7.

29. Citron M, Reynolds J, Kalra J, Kay B, Nathan K, Jaffe N, et al. Pharmacokinetic comparison of intranasal, oral, and intramuscular metoclopramide in healthy volunteers. Cancer treatment reports. 1987;71(3):317–9.

30. Do Nascimento D, de Moraes R, Camarão G, Bezerra F, de Moraes M, de Moraes M. Effect of dimeticone and pepsin on the bioavailability of metoclopramide in healthy volunteers. Drug research. 2014:537–40.

31. Fairhead A, Brooks S, Butterworth K, Mangham B. An automated high-performance liquid chromatographic trace enrichment method for the determination of metoclopramide in serum and its application to a bioequivalence human volunteer study. Food and chemical toxicology. 1989;27(5):341–5.

32. Fass R, Pieniaszek H, Thompson J. Pharmacokinetic comparison of orally-disintegrating metoclopramide with conventional metoclopramide tablet formulation in healthy volunteers. Alimentary pharmacology & therapeutics. 2009;30(3):301–6.

33. Lamparczyk H, Chmielewska A, Konieczna L, Plenis A, Zarzycki PK. RP-HPLC method with electrochemical detection for the determination of metoclopramide in serum and its use in pharmacokinetic studies. Biomedical Chromatography. 2001;15(8):513–7.

34. Leucuþa A, Vlase L, Farcãu D, Nanulescu M. Pharmacokinetic interaction study between ranitidine and metoclopramide. Romanian journal of gastroenterology. 2004;13(3):211–4.

35. Ross-Lee L, Eadie M, Hooper W, Bochner F. Single-dose pharmacokinetics of metoclopramide. European Journal of Clinical Pharmacology. 1981;20:465–71.

36. Vachharajani NN, Shyu WC, Barbhaiya RH. Pharmacokinetic interaction between butorphanol nasal spray and oral metoclopramide in healthy women. The Journal of Clinical Pharmacology. 1997;37(10):979–85.

37. Vlase L, Leucuta A, Farcau D, Nanulescu M. Pharmacokinetic interaction between fluoxetine and metoclopramide in healthy volunteers. Biopharmaceutics & drug disposition. 2006;27(6):285–9.

38. Dallmann A, Pfister M, van den Anker J, Eissing T. Physiologically based pharmacokinetic modeling in pregnancy: a systematic review of published models. Clinical Pharmacology & Therapeutics. 2018;104(6):1110–24.

39. Lau Moon Lin M, Robinson PD, Flank J, Sung L, Dupuis LL. The safety of metoclopramide in children: a systematic review and meta-analysis. Drug safety. 2016;39(7):675–87.

40. Abduljalil K, Pansari A, Jamei M. Prediction of maternal pharmacokinetics using physiologically based pharmacokinetic models: assessing the impact of the longitudinal changes in the activity of CYP1A2, CYP2D6 and CYP3A4 enzymes during pregnancy. Journal of pharmacokinetics and pharmacodynamics. 2020;47(4):361–83.

41. Abduljalil K, Pansari A, Ning J, Jamei M. Prediction of maternal and fetal acyclovir, emtricitabine, lamivudine, and metformin concentrations during pregnancy using a physiologically based pharmacokinetic modeling approach. Clinical Pharmacokinetics. 2022;61(5):725–48.

42. Saunders NR, Liddelow SA, Dziegielewska KM. Barrier mechanisms in the developing brain. Frontiers in pharmacology. 2012;3:46.

43. Saunders N, Habgood M, Dziegielewska K. Barrier mechanisms in the brain, II. Immature brain. Clinical and Experimental Pharmacology and Physiology. 1999;26(2):85–91.

44. Dermietzel R, Krause D, Kremer M, Wang C, Stevenson B. Pattern of glucose transporter (Glut 1) expression in embryonic brains is related to maturation of bood-brain barrier tightness. Developmental dynamics. 1992;193(2):152–63.

45. Cornford EM, Braun LD, Oldendorf WH. Developmental modulations of blood-brain barrier permeability as an indicator of changing nutritional requirements in the brain. Pediatric research. 1982;16(4):324–8.

